# Multi-Modality Machine Learning Models to Predict Stroke and Atrial Fibrillation in Patients with Heart Failure

**DOI:** 10.1101/2023.11.15.23298562

**Authors:** Jiandong Zhou, Lakshmi Murugappan, Lei Lu, Oscar Hou In Chou, Bernard Man Yung Cheung, Gary Tse, Tingting Zhu

## Abstract

**Introduction:** Atrial fibrillation (AF) and stroke are leading causes of death of heart failure patients. Several ML models have been built using electrocardiography (ECG)-only data, or lab test data or health record data to predict these outcomes. However, a multi-modal approach using wearable ECG data integrated with lab tests and electronic health records (EHRs) data has not been developed.

**Objective:** The aim of this study was to apply machine learning techniques to predict stroke and AF amongst heart failure patients from a multi-modal dataset.

**Methods:** This study analysed hospitalised patients with heart failure in Hong Kong between 1 January 2010 and 31 December 2016, with the last follow-up of 31 December 2019. The primary outcomes were AF and stroke. The secondary outcomes were all-cause and cardiovascular mortality. ECG-only, non-ECG-only and multimodal models were built to assess feature importance. Four machine learning classifiers and seven performance measures were used to evaluate the performance.

**Results:** There are in total 2,868 subjects with heart failure upon admission, among them 1,150 (40.10%) had new onset AF, 668 (23.29%) had new onset stroke/TIA. It was found that accurate and sensitive machine learning models can be created to predict stroke and AF from multimodal data. XGBoost, which was the best algorithm tested, achieved a mean (over 10 iterations) accuracy, AUROC, AUPRC, positive predictive value and negative predictive value of 0.89, 0.80, 0.74, 0.99 and 0.88, respectively, for stroke and 0.78, 0.82, 0.77, 0.77 and 0.79, respectively, for AF. The predictive models, built using multimodal data, were easy to use and had high accuracy.

**Conclusion:** Multi-modal machine learning models could be used to predict future stroke and AF occurrences in patients hospitalised for heart failure.

## I. Introduction

Heart Failure (HF) is a clinical condition caused by either structural or functional defects in the myocardium resulting in impairment of ventricular filling or ejection of blood [1]. With millions of individuals affected worldwide, HF poses a pervasive and often devastating challenge within the realm of cardiovascular health [2]. In Hong Kong, HF contributed to 59 episodes of hospitalization per 100,000 population each year [3]. Because HF and atrial fibrillation (AF) are closely inter-related with similar risk factors and shared pathophysiology, they frequently coexist, while AF stands as the most prevalent global arrhythmia and is currently on the rise [4], [5]. Patients with concomitant HF and AF suffer from even worse symptoms and poorer prognosis than those with either of these conditions alone [6]. Patients with AF experience HF have a risk of mortality that is approximately two to threefold higher than that of those without AF [7]. In addition, HF and AF together increase the risks of stroke or transient ischaemic attack, which is the second most common cause of death and the leading cause of disability globally [8]–[10].

The relationships between HF, AF, and stroke are complex, which together complicate the treating outcomes [11]. AF is well known as an independent risk factor for ischaemic stroke, and previous studies showed that this risk is increased by a factor of five in patients with AF [12]. However, less is known about the occurrence of stroke in patients with HF, especially those without AF [13]. Although HF leads to an increase in stroke severity, there is no difference in stroke risk between different HF subtypes [9]. In light of these complexities, it is important to investigate the intricate relationships and the healthcare outcomes for patients with these conditions. This allows identifying high-risk patients necessitating preventive measures, and enabling early detection and treatment of these conditions to prevent further health deterioration. For example, HF and AF are often unrecognised and untreated, because they are frequently asymptomatic or minimally symptomatic; thus, methods to screen for and identify undetected HF and AF are of significant interest to ultimately prevent strokes [14], [15].

Machine learning has seen a growing application in the analysis of electronic health record (EHR) data and electrocardiogram (ECG) data for predicting outcomes related to these conditions. For HF, an artificial intelligence-based clinical decision support system (AI-CDSS) was developed to distinguish HF patients with reduced ejection fraction, mid-range ejection fraction, and preserved ejection fraction [16]; Additionally, a decision tree model was employed to assess mortality risk in both hospitalized and ambulatory HF patients [17]. A convolutional neural network was developed to predict the five-year incident AF risk using 12-lead ECGs [18]. Several studies have also explored health outcomes associated with the coexistence of HF, AF, and stroke. For instance, a deep learning model was designed to predict new-onset AF and identify individuals at risk of AF-related stroke [15]. Previously, it was demonstrated that the ML algorithm had better accuracy in predicting mortality and hospitalization in the setting of acute HF [19]. These studies underscore the potential of machine learning models in improving health outcomes through risk assessment and prediction [20].

This study aims to employ advanced computational methods, specifically utilizing machine learning techniques, to predict the occurrence of stroke/TIA and AF in patients diagnosed with HF.

## II. Results

### A. Basic characteristics

There were in total 2,868 subjects with heart failure upon admission, among them 1,150 (40.10%) had new onset AF, 668 (23.29%) had new onset stroke/TIA, 604 (21.06%) passed away with cardiovascular diseases, and 2,084 (72.66%) passed away with all-causes. The prevalence of the primary and the secondary outcomes are detailed in Table III. A summary of baseline and clinical characteristics in HF patients, including patients with new-onset AF and stroke/TIA in patients can be found in Table IV. The incidence of the adverse outcomes was also calculated (Table V).

**TABLE I:** The multimodal model discussed in this report (right-most column), is compared to other XGBoost models- the combination of multimodal and ECG-only (simple) data as well as individual and multilabel classifiers. All models were trained/tested on the same samples in each iteration. Stroke/TIA outcome had the biggest increase in performance with the use of multimodal data, instead of ECG-only data.

| Atrial Fibrillation |  |  |  |  |  |
| --- | --- | --- | --- | --- | --- |
|  | XGB simple | XGB multilabel | TabNet simple | XGB multimodal+label | XGB multimodal |
| Accuracy | 0.7172±0.0184 | 0.7198±0.0124 | 0.6271 | 0.7823±0.0084 | 0.7814±0.0138 |
| AUROC | 0.7381±0.0206 | 0.7459±0.0078 | 0.6946 | 0.8119±0.0077 | 0.8190±0.0206 |
| AUPRC | 0.6728±0.0300 | 0.6680±0.0139 | 0.6352 | 0.7542±0.0102 | 0.7672±0.0286 |
| PPV | 0.6824±0.0265 | 0.7002±0.0269 | 0.5323 | 0.7835±0.0205 | 0.7704±0.0137 |
| NPV | 0.7340±0.0159 | 0.7292±0.0153 | 0.6994 | 0.7820±0.0109 | 0.7871±0.0155 |
| Stroke/TIA |  |  |  |  |  |
|  | XGB simple | XGB multilabel | TabNet simple | XGB multimodal+label | XGB multimodal |
| Accuracy | 0.7453±0.0105 | 0.7629±0.0069 | 0.7735 | 0.9030±0.0059 | 0.8981±0.0079 |
| AUROC | 0.5329±0.0207 | 0.5380±0.0126 | 0.5168 | 0.8038±0.0108 | 0.8028±0.0237 |
| AUPRC | 0.2606±0.0196 | 0.2604±0.0128 | 0.2630 | 0.7534±0.0121 | 0.7449±0.0253 |
| PPV | 0.2785±0.0975 | 0.2192±0.1467 | 1.0000 | 0.9856±0.0163 | 0.9861±0.0143 |
| NPV | 0.7685±0.0047 | 0.7669±0.0065 | 0.7719 | 0.8895±0.0065 | 0.8844±0.0078 |
| CVD Mortality |  |  |  |  |  |
|  | XGB simple | XGB multilabel | TabNet simple | XGB multimodal+label | XGB multimodal |
| Accuracy | 0.7744±0.0061 | 0.7860±0.0065 | 0.7631 | 0.7790±0.0081 | 0.7774±0.0063 |
| AUROC | 0.5911±0.0261 | 0.5952±0.0132 | 0.6026 | 0.5891±0.0169 | 0.5952±0.0369 |
| AUPRC | 0.2770±0.0264 | 0.2713±0.0121 | 0.2705 | 0.2693±0.0155 | 0.2861±0.0301 |
| PPV | 0.3342±0.0619 | 0.3526±0.2641 | 0.2308 | 0.2946±0.0948 | 0.3682±0.0616 |
| NPV | 0.7949±0.0031 | 0.7893±0.0058 | 0.7883 | 0.7912±0.0060 | 0.7963±0.0026 |
| All-Cause Mortality |  |  |  |  |  |
|  | XGB simple | XGB multilabel | TabNet simple | XGB multimodal+label | XGB multimodal |
| Accuracy | 0.7202±0.0152 | 0.7238±0.0081 | 0.7275 | 0.7664±0.0099 | 0.7669±0.0160 |
| AUROC | 0.6358±0.0222 | 0.6388±0.0119 | 0.5983 | 0.7638±0.0158 | 0.7663±0.0239 |
| AUPRC | 0.8025±0.0147 | 0.8107±0.0110 | 0.7813 | 0.8841±0.0105 | 0.8871±0.0120 |
| PPV | 0.7543±0.0087 | 0.7392±0.0111 | 0.7439 | 0.7950±0.0099 | 0.8021±0.0103 |
| NPV | 0.4754±0.0647 | 0.4985±0.0703 | 0.5000 | 0.6246±0.0354 | 0.6115±0.0466 |

**TABLE II:** Hyperparameters for simple multilabel and multimodal+multilabel models-values are for best iteration out of 10. Performance of the ML models in the subsets.

|  | Multilabel | Multilabel+Multimodal |
| --- | --- | --- |
| max_depth | 3 | 4 |
| learning_rate | 0.0285 | 0.1192 |
| subsample | 0.5216 | 0.5938 |
| colsample_bytree | 0.5220 | 0.7011 |
| min_child_weight | 8.7625 | 8.5115 |

Proof of the First Zonklar Equation

**TABLE III:** Number of cases of each outcome in the dataset.

| Outcome | Number of patients | Percentage |
| --- | --- | --- |
| All-cause mortality | 2084 | 72.66% |
| CVD mortality | 604 | 21.06% |
| Atrial fibrillation | 1150 | 40.10% |
| Stroke/TIA | 668 | 23.29% |

**TABLE IV:**
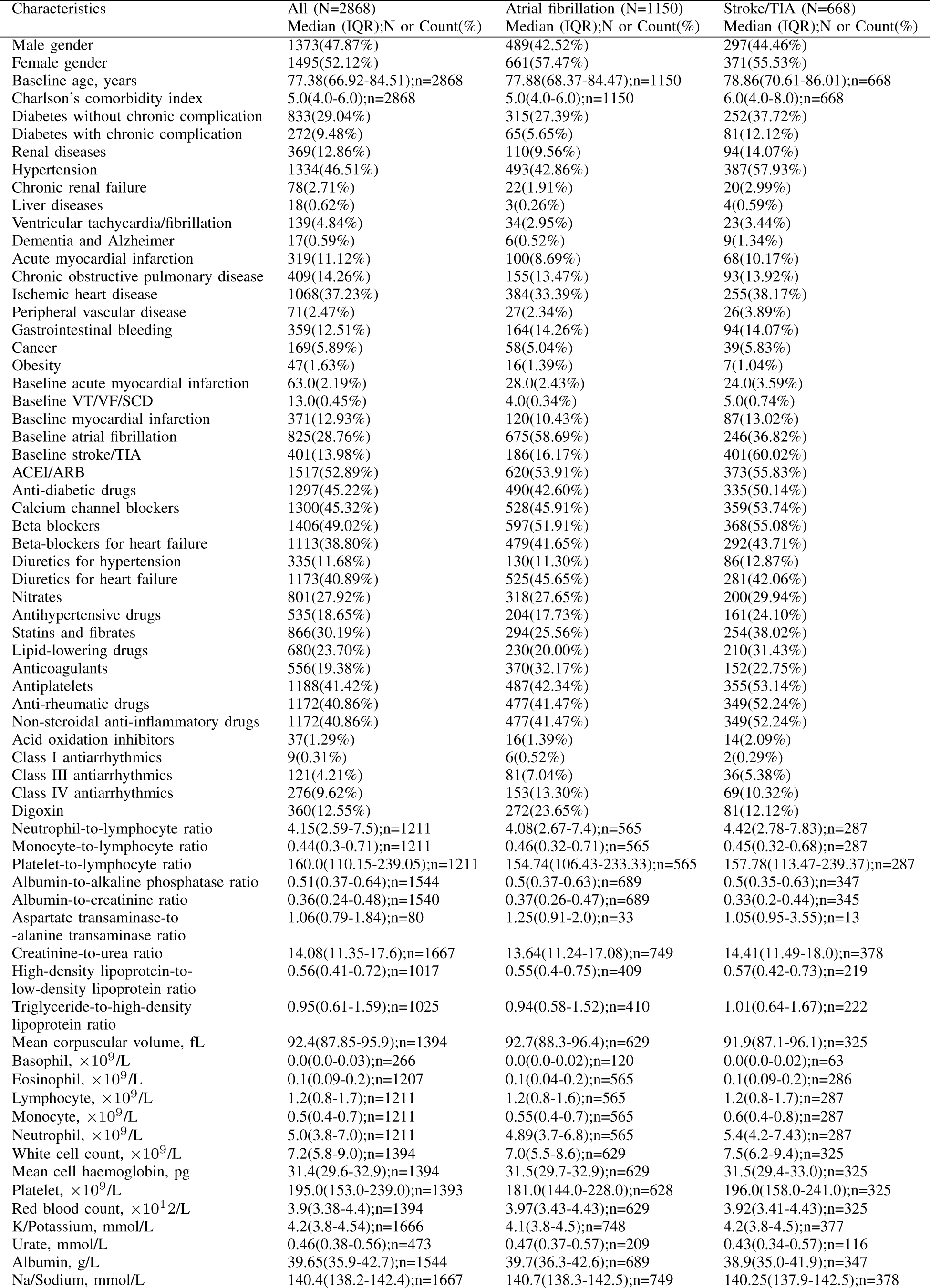
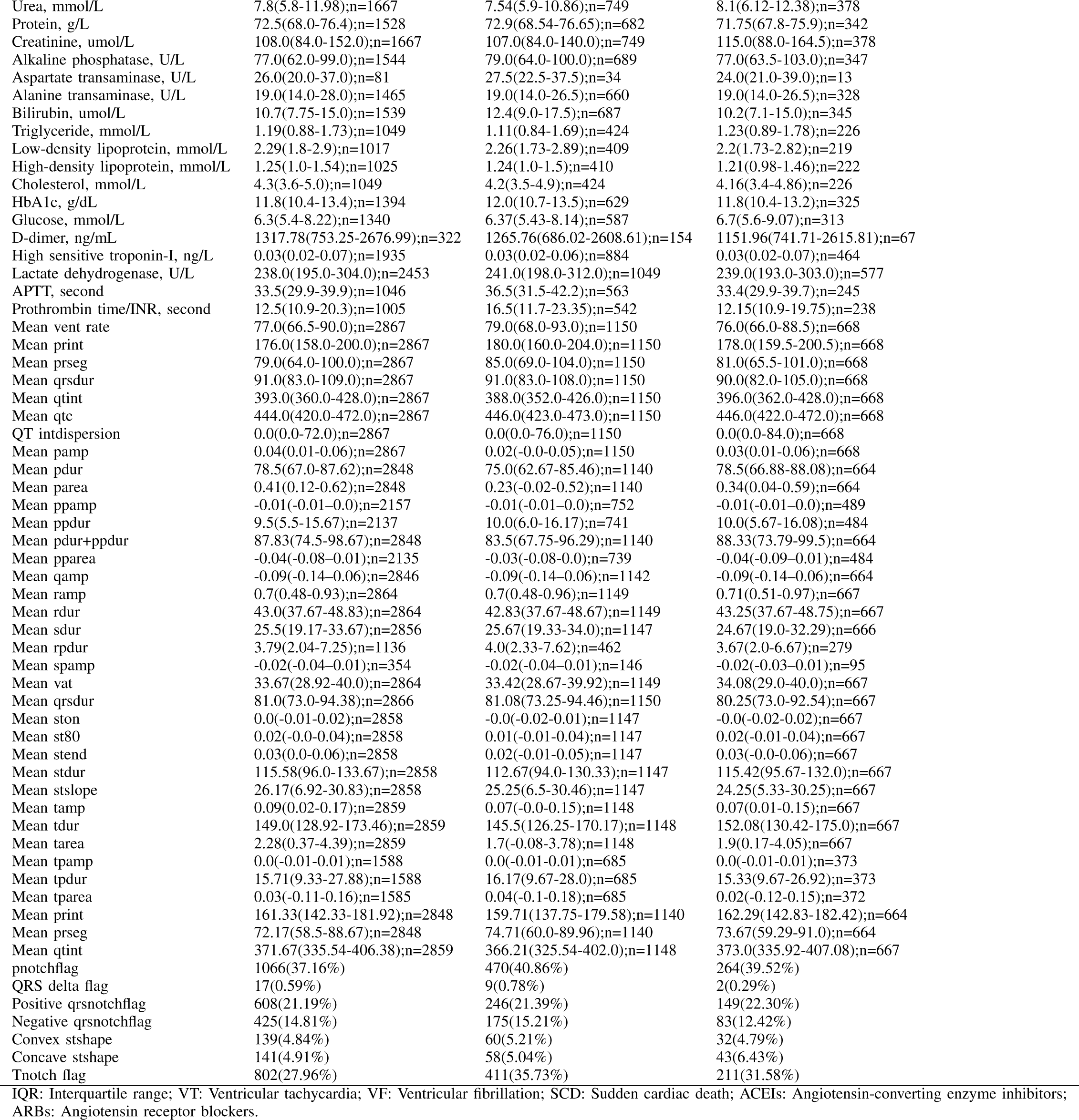
Summary of baseline and clinical characteristics in heart failure patients with new onset AF and stroke/TIA (transient ischemic attack) in patients.

**TABLE V:** Summary of person-year calculations and annualized incidence ratio of new onset AF, new onset stroke/TIA, and mortality risk events in patients with heart failure.

| New onset atrial fibrillation |  |  |  |  |  |
| --- | --- | --- | --- | --- | --- |
| Cohort, year | Number of person-years | Number of events | Incidence rate per 1000 patients | Lower bound | Upper bound |
| 0-1 | 2506.8 | 205 | 81.8 | 71.3 | 93.8 |
| 1-2 | 2141.3 | 122 | 57 | 47.7 | 68 |
| 2-3 | 1834.2 | 124 | 67.6 | 56.7 | 80.6 |
| 3-4 | 1573.5 | 118 | 75 | 62.6 | 89.8 |
| 4-5 | 1352.9 | 109 | 80.6 | 66.8 | 97.2 |
| 5-6 | 1157 | 88 | 76.1 | 61.7 | 93.7 |
| 6-7 | 983.2 | 78 | 79.3 | 63.5 | 99 |
| 7-8 | 822.1 | 75 | 91.2 | 72.8 | 114.4 |
| 8-9 | 662.8 | 68 | 102.6 | 80.9 | 130.1 |
| >9 | 500.6 | 163 | 325.6 | 279.3 | 379.6 |
| Total | 13534.5 | 1150 | 85 | 80.2 | 90 |
| New onset stroke/TIA |  |  |  |  |  |
| Cohort, year | Number of person-years | Number of events | Incidence rate per 1000 patients | Lower bound | Upper bound |
| 0-1 | 2506.8 | 188 | 75 | 65 | 86.5 |
| 1-2 | 2133.5 | 95 | 44.5 | 36.4 | 54.4 |
| 2-3 | 1836.7 | 75 | 40.8 | 32.6 | 51.2 |
| 3-4 | 1582.7 | 69 | 43.6 | 34.4 | 55.2 |
| 4-5 | 1370.5 | 51 | 37.2 | 28.3 | 49 |
| 5-6 | 1187.1 | 33 | 27.8 | 19.8 | 39.1 |
| 6-7 | 1026.6 | 30 | 29.2 | 20.4 | 41.8 |
| 7-8 | 882 | 28 | 31.7 | 21.9 | 46 |
| 8-9 | 740.2 | 38 | 51.3 | 37.4 | 70.6 |
| >9 | 571.1 | 61 | 106.8 | 83.1 | 137.3 |
| Total | 13837 | 668 | 48.3 | 44.8 | 52.1 |
| Cardiovascular mortality |  |  |  |  |  |
| Cohort, year | Number of person-years | Number of events | Incidence rate per 1000 patients | Lower bound | Upper bound |
| 0-1 | 2538.3 | 168 | 66.2 | 56.9 | 77 |
| 1-2 | 2203.6 | 83 | 37.7 | 30.4 | 46.7 |
| 2-3 | 1916.8 | 77 | 40.2 | 32.1 | 50.2 |
| 3-4 | 1668.5 | 59 | 35.4 | 27.4 | 45.6 |
| 4-5 | 1454.1 | 66 | 45.4 | 35.7 | 57.8 |
| 5-6 | 1264 | 31 | 24.5 | 17.2 | 34.9 |
| 6-7 | 1101.3 | 29 | 26.3 | 18.3 | 37.9 |
| 7-8 | 944.6 | 25 | 26.5 | 17.9 | 39.2 |
| 8-9 | 797.1 | 22 | 27.6 | 18.2 | 41.9 |
| >9 | 619.2 | 44 | 71.1 | 52.9 | 95.5 |
| Total | 14507.7 | 604 | 41.6 | 38.4 | 45.1 |
| All-cause mortality |  |  |  |  |  |
| Cohort, year | Number of person-years | Number of events | Incidence rate per 1000 patients | Lower bound | Upper bound |
| 0-1 | 2538.3 | 513 | 202.1 | 185.3 | 220.4 |
| 1-2 | 2203.6 | 280 | 127.1 | 113 | 142.9 |
| 2-3 | 1916.8 | 269 | 140.3 | 124.5 | 158.2 |
| 3-4 | 1668.5 | 193 | 115.7 | 100.5 | 133.2 |
| 4-5 | 1454.1 | 201 | 138.2 | 120.4 | 158.7 |
| 5-6 | 1264 | 146 | 115.5 | 98.2 | 135.8 |
| 6-7 | 1101.3 | 142 | 128.9 | 109.4 | 152 |
| 7-8 | 944.6 | 105 | 111.2 | 91.8 | 134.6 |
| 8-9 | 797.1 | 77 | 96.6 | 77.3 | 120.8 |
| >9 | 619.2 | 158 | 255.1 | 218.3 | 298.2 |
| Total | 14507.7 | 2084 | 143.6 | 137.6 | 149.9 |

The cumulative incidence curves illustrating primary and secondary outcomes stratified by age (Figure 7), sex (Figure 6), and prior major adverse cardiovascular events (MACE) (Figure 8). The analysis revealed an association between age at admission and increased Charlson’s standard comorbidity index with increased risks of AF, stroke/TIA, and increased mortality risks in patients. This association were substantiated by the conditional margin effects analysis (Figure 9 and Figure 10.

### B. ML model prediction performance

The establishment of train/test sets, illustrated in Figure 1, facilitated rigorous testing. The multi-modal model’s outcomes, presented in Table I alongside alternative iterations of the machine learning model for comparative analysis, were derived from 10 distinct train/test sets (80/20 split) to ensure robustness. Remarkably, the results underscored the superior performance achieved with multi-modal data compared to ECG-only data. Moreover, individual classifiers exhibited marginally enhanced performance compared to an XGBoost multilabel classifier, substantiating the efficacy of the proposed multi-modal approach in predictive modeling.

**Fig. 1:**
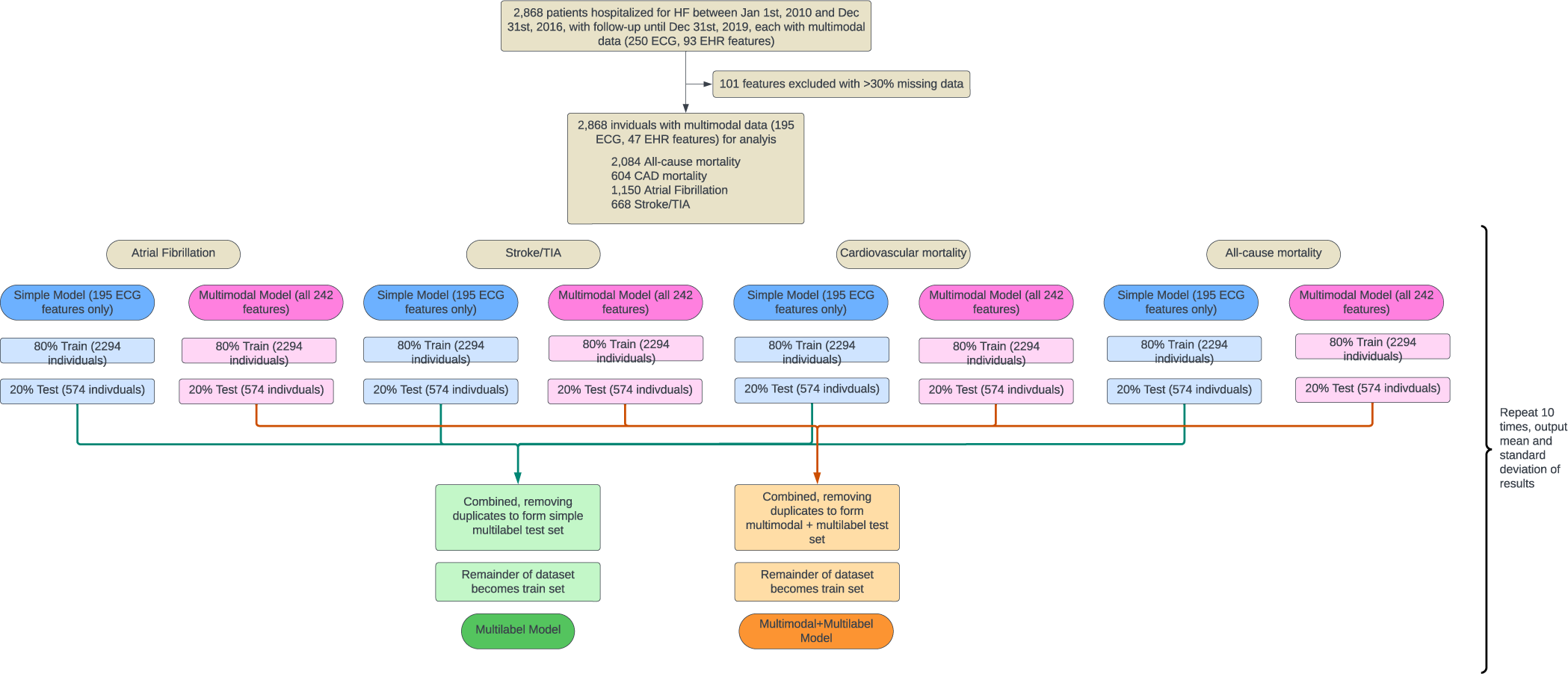
Procedures of constructing training and testing dataset for comparing the various models- ECG-only, multimodal and corresponding multilabel models.

The predictive performance for the stroke/TIA was suboptimal when relying solely on electrocardiogram (ECG) data, with an area under the precision-recall curve (AUPRC) of 0.2606±0.00196. However, an enhancement was observed with the incorporation of electronic health record (EHR) data, resulting in an improved AUPRC of 0.7449±0.0253. AF demonstrated commendable predictive performance independently, registering an AUPRC of 0.7672±0.0286. Assessing positive predictive value (PPV) and negative predictive value (NPV) metrics, both Stroke/TIA (PPV of 0.9861±0.0143 and NPV of 0.8844±0.0078) and AF (PPV of 0.7704±0.0137 and NPV of 0.7871±0.0155) yielded balanced outcomes, with the former exhibiting superior performance. This highlights the pivotal role of integrating EHR data in augmenting the predictive capacity for Stroke/TIA; meanwhile, AF maintains robust predictive capabiliy.

### C. Significant predictors of outcomes

Significant predictors for the individual classifiers were elucidated (Figure 2). In predicting future stroke events, the baseline stroke/TIA emerged as the most significant feature, followed by Charlson’s comorbidity index (CCI). Notably, ECG features, such as max-min ST duration, SD duration, and ST slope, held their positions in the hierarchy of importance, although their contributions were less than a quarter of that to baseline stroke/TIA. Moreover, calcium channel blockers ranked fourth in importance, elucidating the enhancement observed in model performance when non-ECG information was incorporated.

**Fig. 2:**
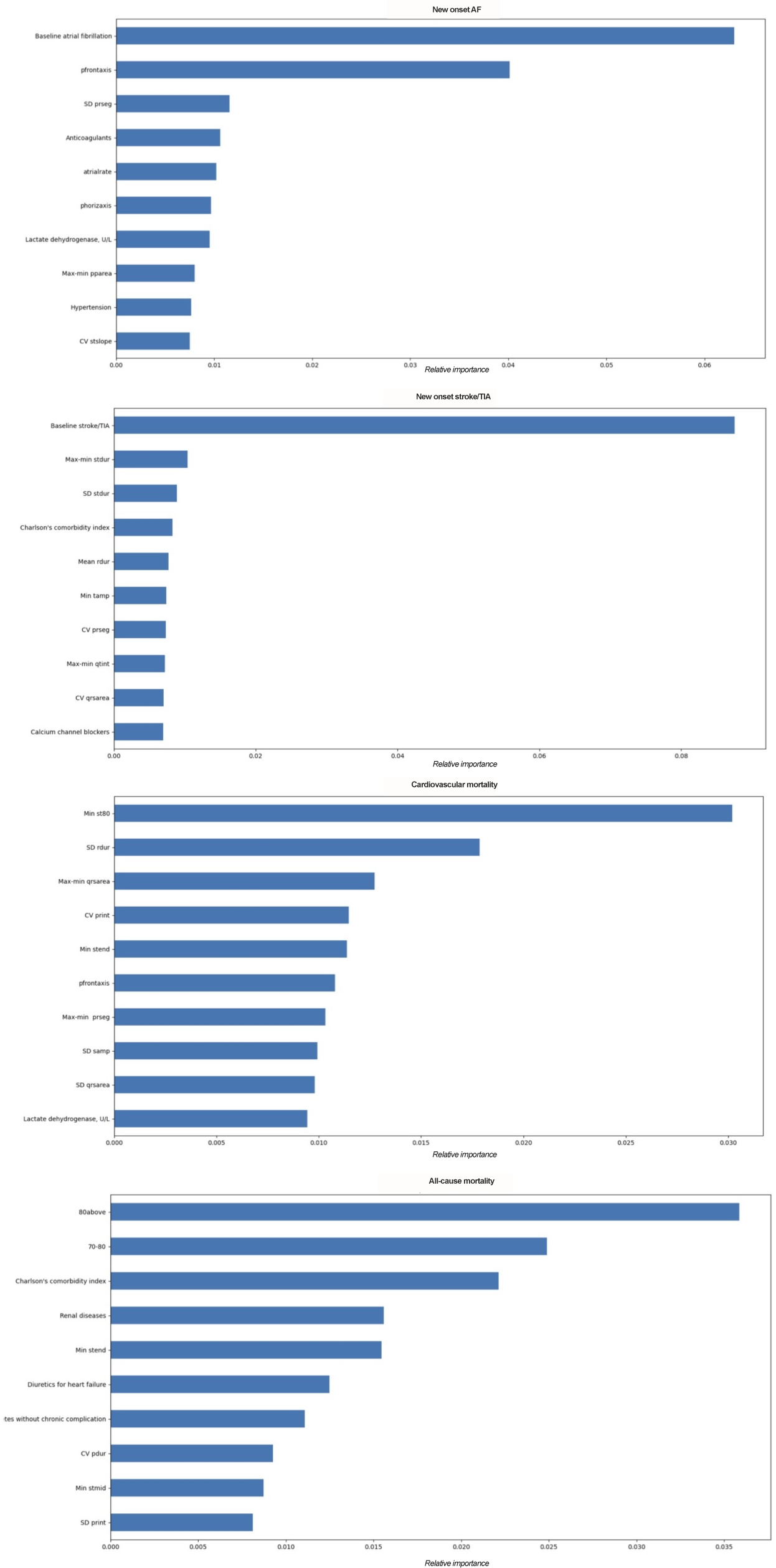
Top 10 important feature rankings to predict new onset AF, new onset stroke/TIA, cardiovascular mortality, and all-cause mortality in patients with heart failure. (The relative importance of each variable in each prediction model was obtained by averaging over 10 iterations.)

Regarding AF, history of prior AF retained its significance as the foremost predictor for recurrence. The second most influential predictor for AF occurrence was ’P Front Axis.’ Subsequently, ECG features assumed pivotal roles, with the SD of the PR segment, atrial rate, and coefficient of variation (CV) of the ST duration emerging as consequential contributors. Additionally, the administration of anticoagulants and the level of lactate dehydrogenase were also identified as critical determinants influencing the risk of AF. This identification of predictive factors not only reaffirmed the importance of historical AF data.

Regarding cardiovascular-associated mortality, a predominant share of predictors originated from ECG data, (Figure 2). Conversely, regarding all-cause mortality, age emerged as the most influential factor, underscoring its intrinsic association with mortality. Additionally, CCI was a significant predictor. Sequentially, the use of diuretics or a diagnosis of heart failure also predicted mortality, preceding the SD of the PR interval. This not only reaffirmed the role of ECG-derived metrics in predicting cardiovascular-associated mortality. It also emphasizes the multifactorial nature of all-cause mortality, wherein age, comorbidity indices, and medications play a significant roles in prognostication.

### D. Model performance evaluation

The evaluation of model performance across diverse patient subgroups is presented in Table VII. Notably, the results underscored the model’s robustness, demonstrating satisfactory performance irrespective of sex, baseline age, and CCI. It is important to note that the subgroup with the least optimal performance manifested in individuals with a CCI of 0-1, with a PPV of 0.5000±0.0024 and a NPV of 0.9203±0.0121. This deviation was attributed to the constraint of a relatively small sample size, encompassing 116 patients within this subgroup. Furthermore, the model exhibited consistent performance for both primary and secondary outcomes across various durations of follow-up, as evidenced by the time-dependent area under the receiver-operating characteristic curve (AUROC) and Harrell’s C-index (Figure 3 and 11).

**Fig. 3:**
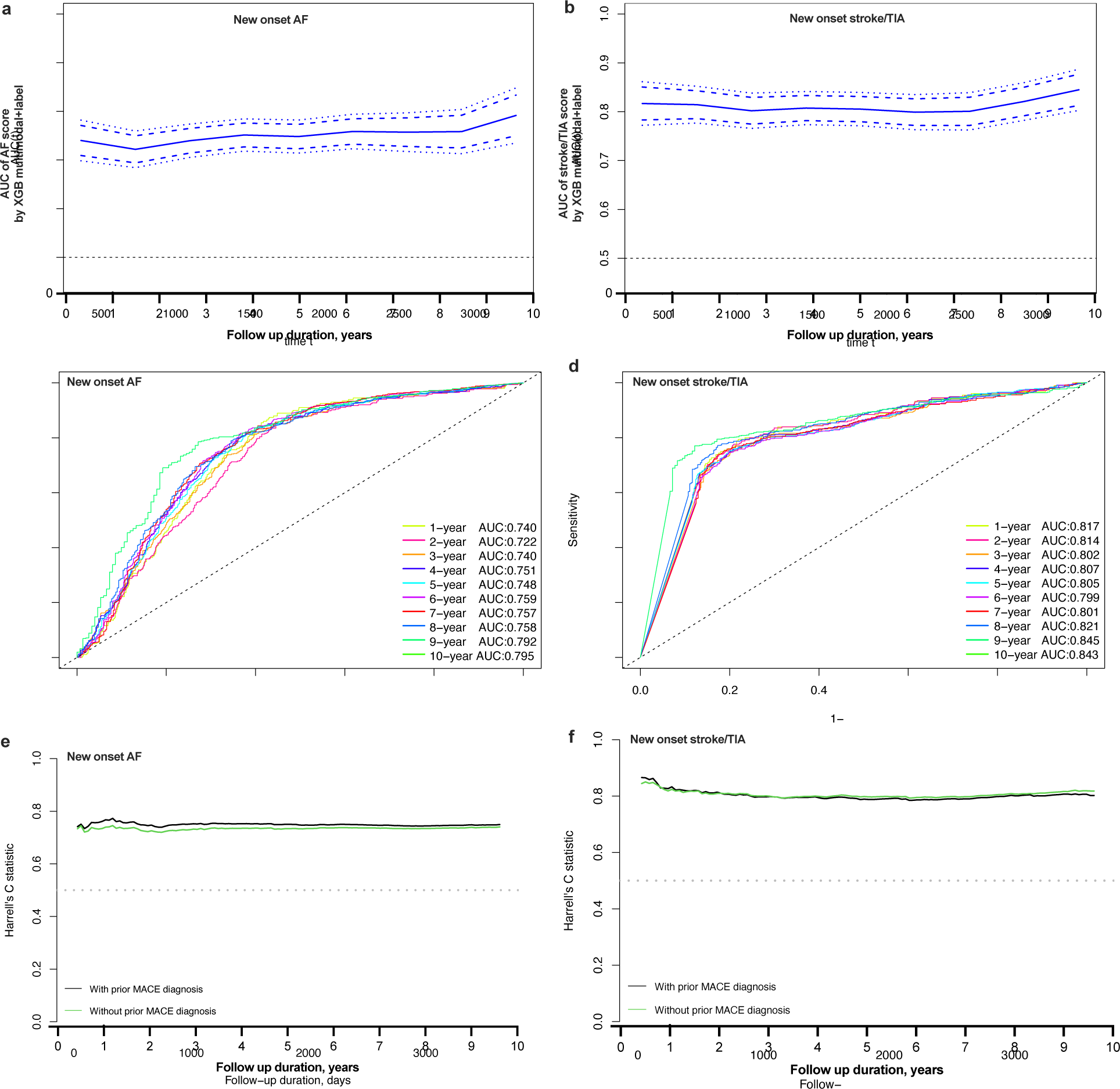
Time-dependent AUROC and Harrell’s C-index of machine learning-based *in-silico* markers to predict new onset AF and new onset stroke/TIA in HF patients. AUC: Area under the receiver operating characteristic curve. AF: Atrial fibrillation. TIA: transient ischemic attack. **a-b**. AUC with 95% confidence interval to predict AF and Stroke/TIA in HF patients with the developed *in-silico* marker by XGB multimodal+label model. **c-d**. Prediction performance measure by AUC to predict new onset AF and new onset stroke/TIA in HF patients with different follow-up duration since admission. **e-f**. Time-dependent C-index of the developed *in-silico* marker to predict new onset AF and new onset stroke/TIA in HF patients with different follow-up duration since admission.

**TABLE VI:** Hyperparameters for simple (ECG-only) and multimodal models- values are for best iteration out of 10.

|  | Simple |  |  |  | Multimodal |  |  |  |
| --- | --- | --- | --- | --- | --- | --- | --- | --- |
|  | All-cause | CVD | AF | Stroke/TIA | All-cause | CVD | AF | Stroke |
| max_depth | 2.0000 | 6.0860 | 3.1011 | 4.1678 | 9.0494 | 7.3671 | 10.8917 | 7.1051 |
| gamma | 3.6418 | 6.1986 | 6.8234 | 7.6980 | 7.3750 | 5.9211 | 7.3989 | 9.3467 |
| min_child_weight | 7.2266 | 16.5424 | 5.2645 | 14.6886 | 15.8779 | 19.2871 | 16.9076 | 17.5608 |
| max_delta_step | 8.4494 | 3.7125 | 9.8757 | 9.6197 | 4.6500 | 8.8195 | 9.0832 | 2.5936 |
| subsample | 0.6546 | 0.5463 | 0.5796 | 0.6198 | 0.8709 | 0.9454 | 0.9112 | 0.4508 |
| colsample_bytree | 0.7818 | 0.5250 | 0.7837 | 0.5102 | 0.8474 | 0.4000 | 0.8894 | 0.6515 |

**TABLE VII:** Model prediction performance on different subgroups of patients.

|  | Gender |  | Age |  | Charlson's Comorbidity Index |  |  |
| --- | --- | --- | --- | --- | --- | --- | --- |
|  | Male | Female | <65 yrs | ≥ 65 yrs | 0-1 | 2-3 | 4+ |
| No. patients | 1373 | 1495 | 625 | 2243 | 116 | 525 | 2227 |
| **Atrial Fibrillation** |  |  |  |  |  |  |  |
| Accuracy | 0.7818±0.0206 | 0.7632±0.0187 | 0.7616±0.0292 | 0.7773±0.0118 | 0.7250±0.0837 | 0.7133±0.0414 | 0.7832±0.0199 |
| AUROC | 0.8073±0.0188 | 0.8177±0.0230 | 0.7827±0.0330 | 0.8287±0.0210 | 0.6647±0.1489 | 0.7680±0.0375 | 0.8257±0.0216 |
| AUPRC | 0.7082±0.0330 | 0.7841±0.0393 | 0.6981±0.0499 | 0.7874±0.0279 | 0.5257±0.1416 | 0.7195±0.0495 | 0.7707±0.0328 |
| PPV | 0.7364±0.0430 | 0.7609±0.0269 | 0.7015±0.0600 | 0.7771±0.0220 | 0.5333±0.2176 | 0.6942±0.0651 | 0.7720±0.0292 |
| NPV | 0.8023±0.0191 | 0.7655±0.0177 | 0.7855±0.0284 | 0.7780±0.0112 | 0.7761±0.0609 | 0.7268±0.0364 | 0.7899±0.0207 |
| **Stroke/TIA** |  |  |  |  |  |  |  |
| Accuracy | 0.9040±0.0060 | 0.9030±0.0148 | 0.9008±0.0176 | 0.9013±0.0128 | 0.9167±0.0186 | 0.8771±0.0090 | 0.9070±0.0133 |
| AUROC | 0.7676±0.0223 | 0.8339±0.0263 | 0.6846±0.0580 | 0.8231±0.0347 | 0.6682±0.1855 | 0.5900±0.1013 | 0.8291±0.0365 |
| AUPRC | 0.7128±0.0184 | 0.7958±0.0337 | 0.5896±0.0730 | 0.7791±0.0383 | 0.3826±0.1875 | 0.3072±0.1015 | 0.8033±0.0389 |
| PPV | 0.9852±0.0126 | 0.9642±0.0364 | 0.9678±0.0700 | 0.9799±0.0135 | 0.5000±0.0024 | 0.7600±0.2268 | 0.9870±0.0097 |
| NPV | 0.8928±0.0060 | 0.8916±0.0144 | 0.8948±0.0140 | 0.8871±0.0133 | 0.9203±0.0121 | 0.8818±0.0070 | 0.8903±0.0142 |
| **CAD Mortality** |  |  |  |  |  |  |  |
| Accuracy | 0.7807±0.0098 | 0.7528±0.0096 | 0.8216±0.0119 | 0.7639±0.0096 | 0.7958±0.0224 | 0.8067±0.0181 | 0.7574±0.0138 |
| AUROC | 0.5609±0.0326 | 0.6073±0.0469 | 0.6531±0.0487 | 0.5908±0.0305 | 0.6032±0.1168 | 0.6741±0.0479 | 0.5765±0.0241 |
| AUPRC | 0.2224±0.0253 | 0.2923±0.0353 | 0.2934±0.0698 | 0.2884±0.0297 | 0.3500±0.1278 | 0.3202±0.0383 | 0.2653±0.0240 |
| PPV | 0.1806±0.1109 | 0.3088±0.0749 | 0.4025±0.1337 | 0.3406±0.0794 | 0.5278±0.0213 | 0.3976±0.1482 | 0.2850±0.0989 |
| NPV | 0.8068±0.0043 | 0.7819±0.0054 | 0.8442±0.0069 | 0.7855±0.0044 | 0.8119±0.0250 | 0.8315±0.0090 | 0.7833±0.0056 |
| **All-cause Mortality** |  |  |  |  |  |  |  |
| Accuracy | 0.7498±0.0197 | 0.7732±0.0152 | 0.7248±0.0370 | 0.8096±0.0131 | 0.7417±0.0764 | 0.6876±0.0392 | 0.8139±0.0144 |
| AUROC | 0.7454±0.0327 | 0.7806±0.0242 | 0.8048±0.0356 | 0.7288±0.0362 | 0.7882±0.0691 | 0.7485±0.0283 | 0.7023±0.0285 |
| AUPRC | 0.8700±0.0249 | 0.9005±0.0165 | 0.7846±0.0503 | 0.9053±0.0151 | 0.6160±0.1322 | 0.7097±0.0373 | 0.9004±0.0126 |
| PPV | 0.7875±0.0129 | 0.8046±0.0120 | 0.7105±0.0347 | 0.8284±0.0081 | 0.6030±0.2833 | 0.6655±0.0506 | 0.8319±0.0094 |
| NPV | 0.5895±0.0544 | 0.6157±0.0480 | 0.7396±0.0445 | 0.5720±0.0962 | 0.7800±0.0479 | 0.7082±0.0421 | 0.5217±0.1192 |

**TABLE VIII:** Performance results of prediction model using non-ECG data.

| Measure | Atrial Fibrillation | Stroke/TIA | CAD Mortality | All-cause Mortality |
| --- | --- | --- | --- | --- |
| Accuracy | 0.7547 ± 0.0171 | 0.8868 ± 0.0092 | 0.7476 ± 0.0101 | 0.7490 ± 0.0163 |
| AUROC | 0.7900 ± 0.0179 | 0.8128 ± 0.0220 | 0.5318 ± 0.0266 | 0.7370 ± 0.0208 |
| AUPRC | 0.7281 ± 0.0209 | 0.7543 ± 0.0253 | 0.2346 ± 0.0188 | 0.8707 ± 0.0126 |
| PPV | 0.7253 ± 0.0242 | 0.8997 ± 0.0274 | 0.2459 ± 0.0525 | 0.7942 ± 0.0102 |
| NPV | 0.7703 ± 0.0149 | 0.8846 ± 0.0080 | 0.7923 ± 0.0046 | 0.5599 ± 0.0440 |

Figure 5 elucidates the enhancement in model performance as additional features are incorporated. Notably, for the prediction of AF, cardiovascular-associated mortality, and all-cause mortality, a surge in performance was observed with the integration of approximately 10 features. Subsequently, the performance levels reached a plateau, signifying a saturation point in the model’s benefit from additional features. In contrast, the prediction of stroke displayed a distinctive pattern, maintaining a consistent level of performance across all features. This was attributed to the overwhelming significance of ’baseline stroke/TIA’ as the preeminent predictor of future stroke events (Figure 2). To discern the specific impact of non-electrocardiogram (non-ECG) features on model performance, four classifiers were trained exclusively on non-ECG data (Table **??**). Intriguingly, AF prediction exhibited the least improvement with the incorporation of multi-modal data compared to non-ECG-only data, whereas all-cause mortality demonstrated the most substantial enhancement. This analysis not only highlighted the optimum feature threshold for different outcomes.

## III. METHODS

### A. Data and setting

This study was approved by The Joint Chinese University of Hong Kong - New Territories East Cluster Clinical Research Ethics Committee. This was a retrospective, territory-wide cohort study of hospitalized patients with ECG measurements between 1^st^ January 2000 and 31^st^ December 2019 from a single tertiary centre in Hong Kong, China. The patients were identified from the Clinical Data Analysis and Reporting System (CDARS) [21]–[23], a territory-wide database that centralizes patient information.

The baseline characteristics of patients were succinctly summarized utilizing descriptive statistics. Continuous variables were expressed as median [95% confidence interval (CI)/interquartile range] or mean [standard deviation (SD)], while categorical variables were presented as total numbers and percentages. To discern differences between continuous variables, the two-tailed Mann–Whitney U test was employed, and for 2 × 2 contingency data, the two-tailed *χ*^2^ test with Yates’ correction was applied. A *P* value *<* 0.05 was indicated of statistical significance. This methodology ensures a comprehensive and reliable exploration of patient characteristics in the study cohort. All statistical analyses were performed with RStudio (Version: 1.1.456) and Python (Version: 3.6).

The dataset employed in this study encompasses a comprehensive set of information, comprising 250 features derived from a 12-lead electrocardiogram ECG recording, as illustrated in Figure 4. Additionally, EHR data, comprising 93 features, encompasses crucial patient details such as gender, age, medical history, medications, and laboratory test results. The investigation focuses on four distinct outcomes: all-cause mortality, cardiovascular mortality, AF, and stroke/TIA. To contextualize these outcomes, the time elapsed from the ECG recording date was meticulously calculated. This comprehensive dataset amalgamation facilitates a nuanced exploration of the multifaceted relationships between ECG features, EHR information, and diverse clinical outcomes.

**Fig. 4:**
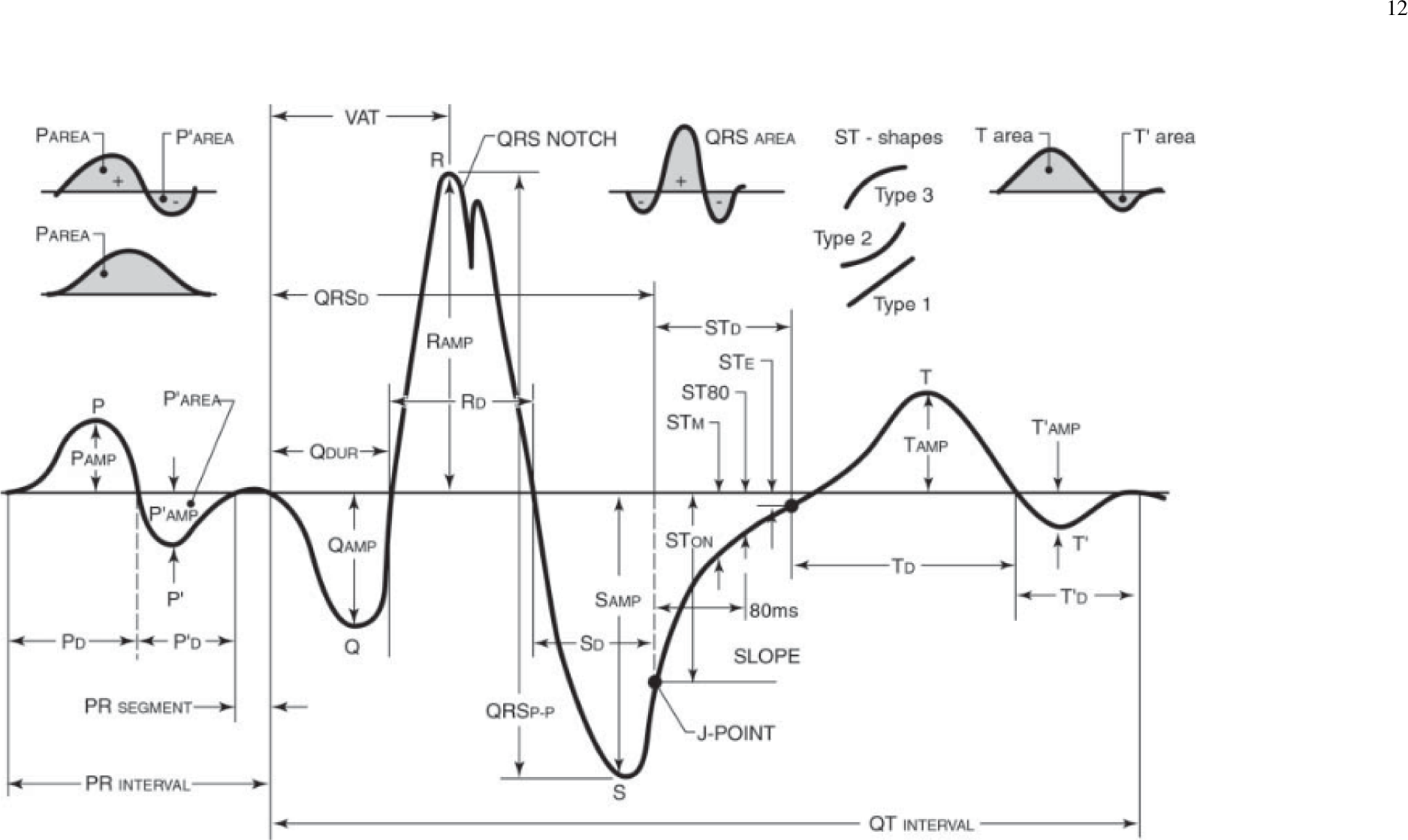
Illustration of 12-lead ECG features. [39]

**Fig. 5:**
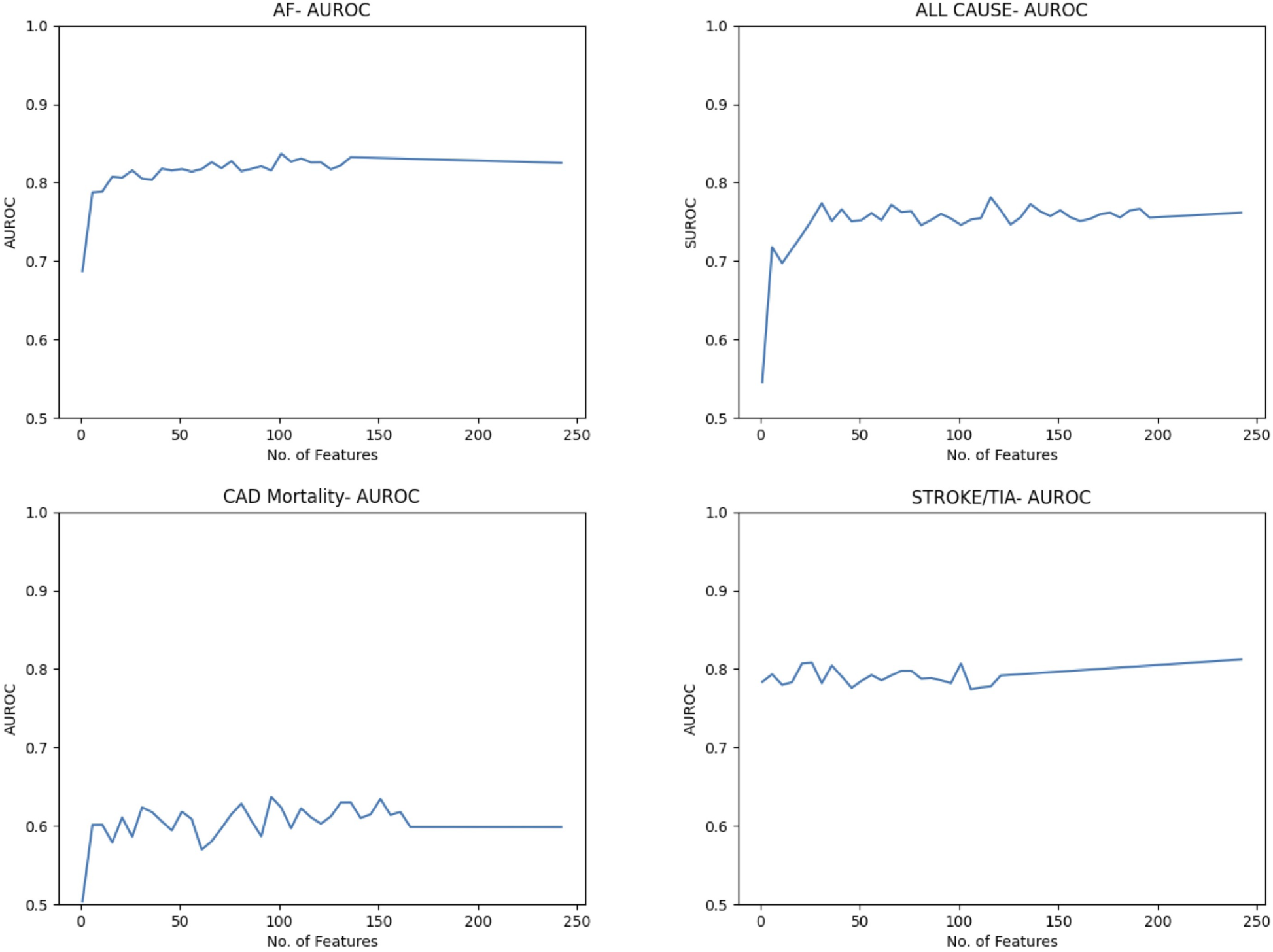
Model performance (AUROC) for each outcome with addition of more features. AUROC values plotted of every 5 features added.

### B. Data pre-processing

It was found that over 50 features had more than 30% of values missing. So the decision was made to eliminate these features entirely reducing the size of the feature vector to 2868x242. Data was then normalised. For this, within each combination, features were scaled using StandardScaler’s ’fit transform’ (for training data) and ’transform’ (for test data) to avoid the model learning the test data, and keeping it unseen.

### C. Prediction task approach

In constructing the multimodal model, four distinct classifiers were developed, each tailored to a specific outcome. The initial step involved partitioning the data into 80% training and 20% testing sets, ensuring a stratified division for each outcome category. Subsequently, a Bayesian parameter search was conducted on the training data to pinpoint the optimal parameters, with a primary focus on maximizing the Area Under the Receiver Operating Characteristic (AUROC) curve. The selected parameters for the multimodal model are delineated in Appendix A. The AUROC metric was deliberately chosen due to the balanced nature of the training samples, ensuring an unbiased assessment of model performance. The parameter search was executed through a meticulous three-fold validation process encompassing 20,000 iterations, contributing to the robustness and reliability of the parameter selection. The parameters to be optimised were:

*•* **min child weight:** The minimum sum of weights of all observations required in a child (derived) node.
*•* **gamma:** The minimum loss reduction required for a node to be split. (I.e. splitting will result in this reduction in loss)
*•* **subsample:** The fraction of observations to be selected for each tree. Selection is done by random sampling.
*•* **colsample bytree:** Similar to max features. Number of columns to be random samples for each tree.
*•* **max depth:** The max depth of the tree, to avoid overfitting. If set to default, the algorithm would aim to continue till all leaf nodes are pure.
*•* **max delta step:** The max step of the update. Positive values ensure a more conservative update- used for imbalanced classes.

Following the identification of optimal parameters through Bayesian parameter search, an XGBoost classifier was trained using the designated parameters on the training dataset. Subsequently, the classifier’s performance was rigorously assessed on the unseen test set, providing a comprehensive evaluation of its generalization capabilities to new and previously unencountered data. This pivotal step ensures a robust understanding of the model’s effectiveness beyond the training context, gauging its potential for real-world applications.

### D. Class-balancing techniques

In addressing the issue of class imbalance, we strategically employed the ’sample weight’ and ’scale pos weight’ parameters within the XGBoost framework, focusing their application solely on the training data. This deliberate choice aimed to rectify disparities among classes, enhancing the classifier’s ability to effectively learn from the training set. The ’sample weight’ parameter assigns varying weights to individual samples, enabling the model to place greater emphasis on underrepresented classes. Simultaneously, the ’scale pos weight’ parameter adjusts the balance between positive and negative class weights, fostering a more nuanced and accurate learning process.

By implementing these parameters exclusively during training, we sought to ensure that the classifier acquired the necessary sensitivity to diverse class distributions without compromising the authenticity of the test data. This approach is crucial for producing a model that not only performs well in a controlled training environment but also generalizes effectively to real-world scenarios. The careful calibration of these parameters contributes to a more robust and adaptive classifier, capable of handling imbalanced class distributions while maintaining relevance to the broader context of diverse outcomes.

To balance classes, the ’sample weight’ and ’scale pos weight’ parameters in XGBoost were used on training data only. This ensured that classes were not too imbalanced and could be used to train the classifier, while also keeping the test data realistic, and symbolic of a real-world distribution of outcomes.

### E. Assessment of performance

The assessment of binary classification performance in this study employed the area under the precision-recall curve (AUPRC), chosen for its resilience to imbalanced test data. Additional metrics, including the AUROC, accuracy, PPV, and NPV, were also considered for a comprehensive evaluation of model prediction performance.

## IV. Discussion

In this investigation, our model demonstrated a significant efficacy in predicting the occurrence of stroke and AF among hospitalized heart failure HF patients. Furthermore, we identified the key predictors contributing significantly to these adverse outcomes. The precision of our predictive model underscores its potential as a valuable tool in clinical settings for risk assessment and prognostication among HF patients. The discernment of influential predictors enhances our understanding of the complex interplay of factors contributing to stroke and AF in this patient cohort, offering valuable insights for tailored intervention strategies and patient management.

### A. Comparison with the previous studies

While the medical treatment for HF has continued to advance, HF remained a major cause of morbidity and mortality worldwide [24]. It was suggested that AF might occur in up to 57% of the patients with HF, and contributed to most of the stroke cases [25]. Furthermore, AF was associated with mortality amongst patients with HF [26]. Meanwhile, HF is a predictor of ischaemic stroke regardless the patients have AF or not. As such, the coexistence of HF and AF further complicated the picture with their potential synergistic effects on stroke development [25].

Machine learning has been used extensively in studying AF, stroke/TIA, or HF. Previously, AI-ECG technology was employed to detect paroxysmal AF during sinus rhythm in patients with cryptogenic stroke [27]. Furthermore, another model integrated carotid ultrasound images and conventional risk factors to stratify the risk of stroke [28]. The multi-modality ML-based models allowed the identification of high stroke-risk patients amongst hospitalised HF patients with an AUROC of 0.8028. A previous literature using UK-Biobank devised six machine learning models to predict ischaemic stroke amongst current AF patients. The best AUROC achieved by the XGBoost model was 0.631, which was reported to have performed better than the CHA2DS2- VASc score based on DeLong’s test (AUROC: 0.611) [29]. Meanwhile, in a study involving 503,842 Chinese adults, the gradient-boosted trees provided the best performance with an AUROC of 0.83. For instance, in a study involving 3,435,224 United States patients, it was reported that the ML-based algorithms outperformed the existing clinical risk scores, and that using the ML models would be more useful than the ’treat all’ strategy [30]. However, no existing literature has incorporated ECG data to identify patients at risk of stroke/TIA amongst hospitalised HF patients.

Regarding AF, a study developed 5 machine learning models for the prediction of new-onset AF amongst ischaemic stroke patients. Their best model, the deep neural network model, had a C index of 0.77 [31]. This was significantly superior to the CHA2DS2-VASc score, Framingham risk score and C2HEST score. In our study, the C index (0.76; Confidence interval: 0.71- 0.29) of the XGBoost multi-modality model was comparable to their deep neural network model, although the targeted groups of patients were different. Meanwhile, another study previously used the component-wise gradient boosting method to identify the extra risk factors for incident AF amongst post-stroke patients using the German health claims data [32]. The AUROC of our study (AUROC: 0.8190) was comparable with that study, which reported an AUROC of 0.829. Furthermore, a study using random survival forests to predict new-onset AF amongst patients with existing cardiovascular disease with cardiovascular magnetic resonance data reported an AUROC of 0.80. The above results demonstrated machine learning approaches were able to identify AF with much higher performance compared to predictions with the conventional AF risk factors.

The multi-modality machine learning-based prediction models allowed the identification of the predictors of stroke. Baseline stroke/TIA was identified to be the most significant predictor. This aligned with the existing literature, in which the risk of stroke recurrence after the first stroke was substantial [33]. Besides, the literature suggested that the ECG features predicting stroke/TIA included QT prolongation, T wave and ST segment abnormalities, atrioventricular block, and prominent U wave [34]. In our model, the ECG features such as max-min ST duration and ST slope, corresponding to the changes of the ST segment reported, predicted the occurrence of stroke. The ST segment slope has been proposed as a predictor of transient myocardial ischaemia or coronary artery disease [35]. While the link between ST slope with ischaemic stroke has been less well reported, we postulated that it might be explained by atherosclerotic changes. Therefore, identifying the subtle changes in automated ECG as such might allow us to predict the risks of stroke/TIA amongst hospitalised HF patients.

For the predictors of AF, the P wave (frontal) axis indicated the anatomical features such as the positioning of the atria and the relative size of the atria. It also reflects the abnormal atrial electrical wavefront propagation in a diseased myocardium. In a retrospective cohort study of US veterans, the P-wave axis was shown to be a significant predictor of AF [5], supporting the findings of the ML model. Meanwhile, atrial high rate episodes, which were defined as an atrial rate limit of ¿175, was reported to be in up to 70% of the AD patients [36]. It was suggested that the atrial high rate episodes were associated with increased risk of AF, increased thromboembolic risk, as well as ischaemic stroke [37]. Those findings provided evidence regarding the robustness of the model in predicting adverse outcomes. For predicting cardiovascular mortality, ECG features remained the most important predictors. This explained the lack of vast improvement in model performance when multi-modal data was used as opposed to ECG-only data. This would also explain the poorer performance in predicting this outcome.

### B. Clinical implications

The integration of diverse data modalities provides a comprehensive perspective, allowing for a more complete understanding of the relationships between these cardiovascular conditions. This approach not only contributes to the advancement of predictive modeling in cardiovascular medicine, but also offers a novel avenue for exploring the interconnected dynamics of HF, AF, and stroke within a single analytical framework. The availability of automated ECG data allows the identification of subtle ECG changes that were not identified in manual extraction. The multi-modality machine learning-based prediction models allow better risk stratification of AF and stroke/TIA of hospitalised HF patients. For example, identifying a patient with a high risk of AF allows justification for using Holter to further monitor the patients to prevent AF-related cardiovascular events [38]. This enables better-personalised survival estimation and timely intervention and management for the patient.

### C. Future work

Future research could look into the time-varying effects of the data after upon baseline event, and use this to make more accurate predictions. This includes the effects of interventions between baseline and adverse events. This might help improve results in predicting AF and stroke/TIA occurrence. Furthermore, the model could be extended, using the ’time-to’ data to predict new onset events after some follow-up duration in a patient. This may improve the care provided to a patient, allowing healthcare professionals to intervene sooner to prevent such events and reduce overall mortality rates. Further, it is of interest to predict MACE recurrence in patients with HF.

### D. Limitations

There were several limitations that should be appreciated in this study. Firstly, given its observational nature, there might be under-coding, coding errors, and missing data resulting in information bias. For instance, baseline tests and information are used to predict mortality without considering time-varying effects in the living period before death and the effects of any interventions. This limits the realistic value of the results.

Secondly, the data was only based on a single locality (Hong Kong), which that the models may require validation in other localities for generalizability. Furthermore, important risk factors for cardiovascular events, such as smoking, alcohol use, and BMI were not readily coded in CDARS. However, we have included multiple comorbidities and laboratory parameters that were closely associated with those missing risk factors. Last but not least, the lack of the echocardiogram data and the NYHA functional class did not allow classification and severity stratification of HF patients. However, as the AUROC and overall accuracy of the outcomes were high, the prediction models still allowed accurate predictions of the adverse outcomes.

## V. Conclusion

The application of multimodal machine learning models, integrating electronic health records and automated ECG data, facilitated the prediction of AF and stroke/TIA among hospitalized HF patients. Notably, the model’s capability to discern subtle ECG changes proved instrumental in identifying HF patients at risks of AF or stroke/TIA. This approach might contribute to more personalized patient care in HF management.

## Funding source

1. L. L was supported by the InnoHK Project at the Hong Kong Centre for Cerebro-cardiovascular Health Engineering (COCHE).
2. T. Z was supported by the Royal Academy of Engineering under the Research Fellowship scheme.

## Data Availability

Data are not available, as the data custodians (the Hospital Authority and the Department of Health of Hong Kong SAR) have not given permission for sharing due to patient confidentiality and privacy concerns. Local academic institutions, government departments, or nongovernmental organizations may apply for access to data through the Hospital Authority's data-sharing portal (https://www3.ha.org.hk/data).

## Acknowledgment

None.

## Conflict of interest

None.

## Guarantor Statement

All authors approved the final version of the manuscript. T. Zhu and G. T is the guarantor of this work and, as such, had full access to all the data in the study and takes responsibility for the integrity of the data and the accuracy of the data analysis.

## Ethical approval statement

This study was approved by the Institutional Review Board of the University of Hong Kong/Hospital Authority Hong Kong West Cluster (HKU/HA HKWC IRB) (UW-20-250), and New Territories East Cluster-Chinese University of Hong Kong (NTEC-UCHK) Clinical Research Ethics Committee (2018.309, 2018.643) and complied with the Declaration of Helsinki.

## Author contributions

Data analysis: J.D. Z., L. M.

Data review: J.D. Z., L. M., O.H.I. C., G. T.

Data acquisition: J.D. Z., O.H.I. C., G. T.

Data interpretation: J.D. Z., O.H.I. C., G. T.

Critical revision of the manuscript: J.D. Z., L. L., L. M., O.H.I. C., B. M. Y. C., G. T., T. Z. Supervision: B. M. Y. C., G. T., T. Z.

Manuscript writing: J.D. Z., L. M., L. L., O.H.I. C., G. T.

Manuscript revision: J.D. Z., L. M., L. L., O.H.I. C., B. M. Y. C., G. T., T. Z.

## Appendix A Proof of the First Zonklar Equation

Appendix one text goes here.

**Fig. 6:**
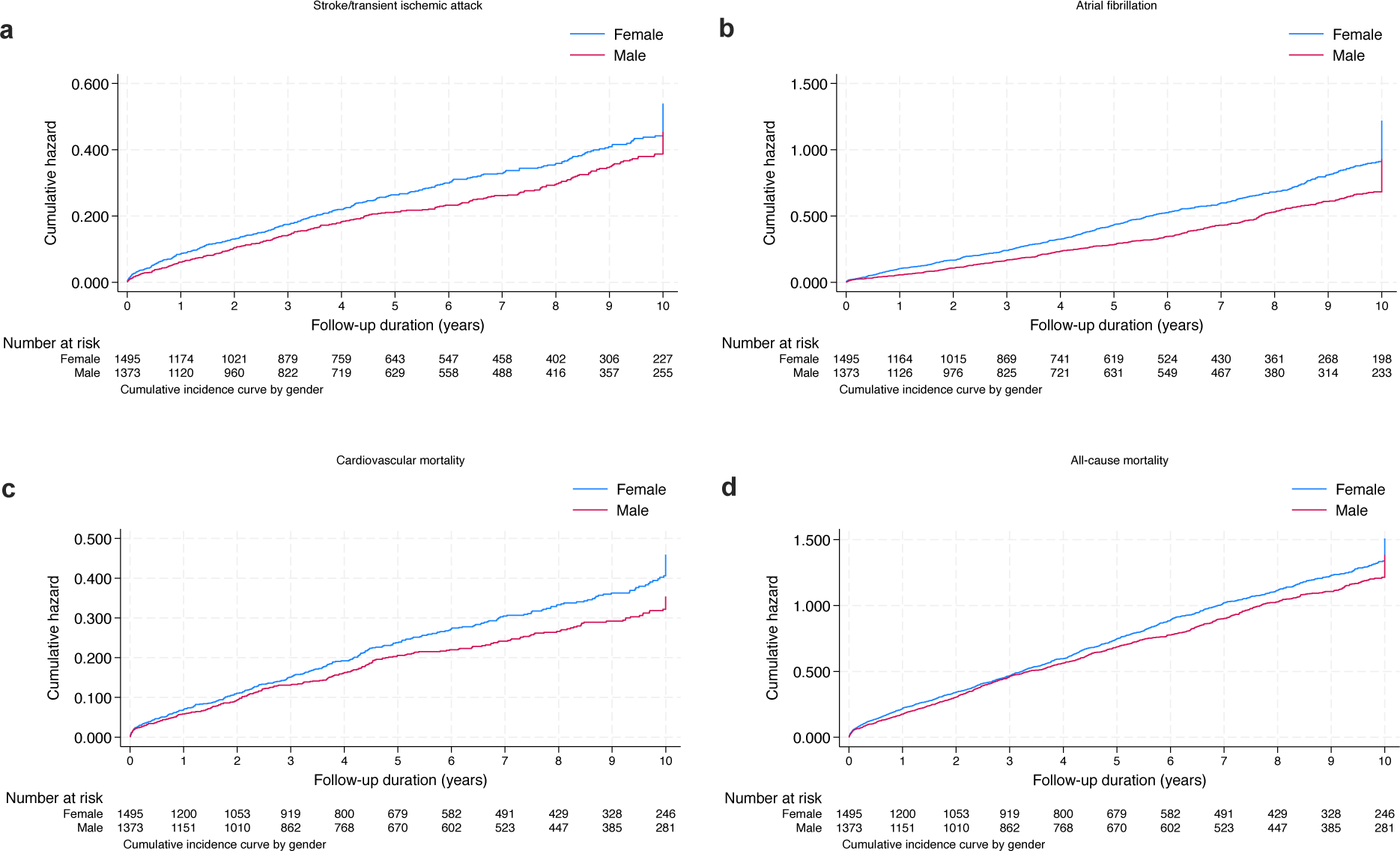
Cumulative incidence curves for primary and secondary outcomes, stratified by sex.

**Fig. 7:**
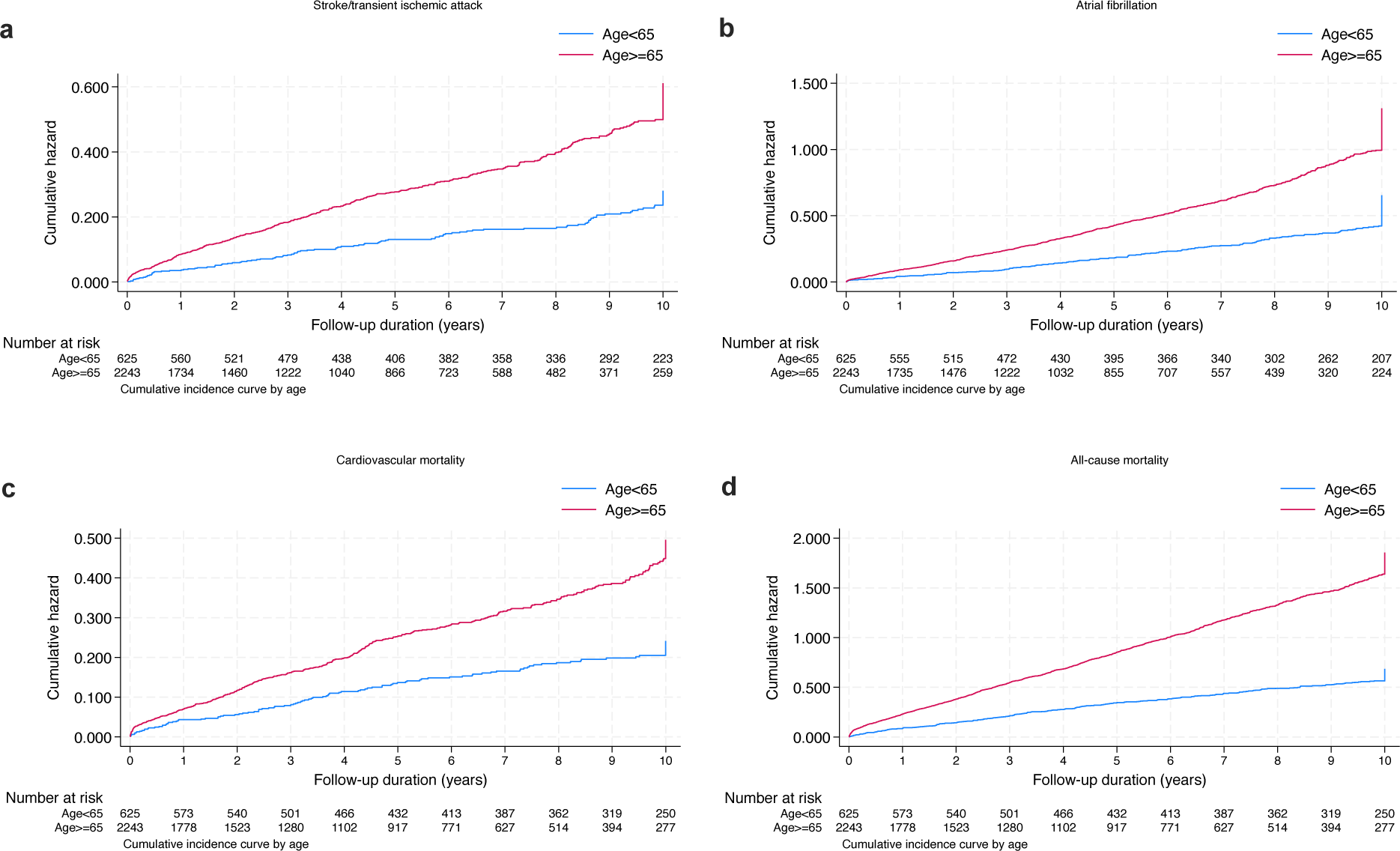
Cumulative incidence curves for primary and secondary outcomes, stratified by age at admission.

**Fig. 8:**
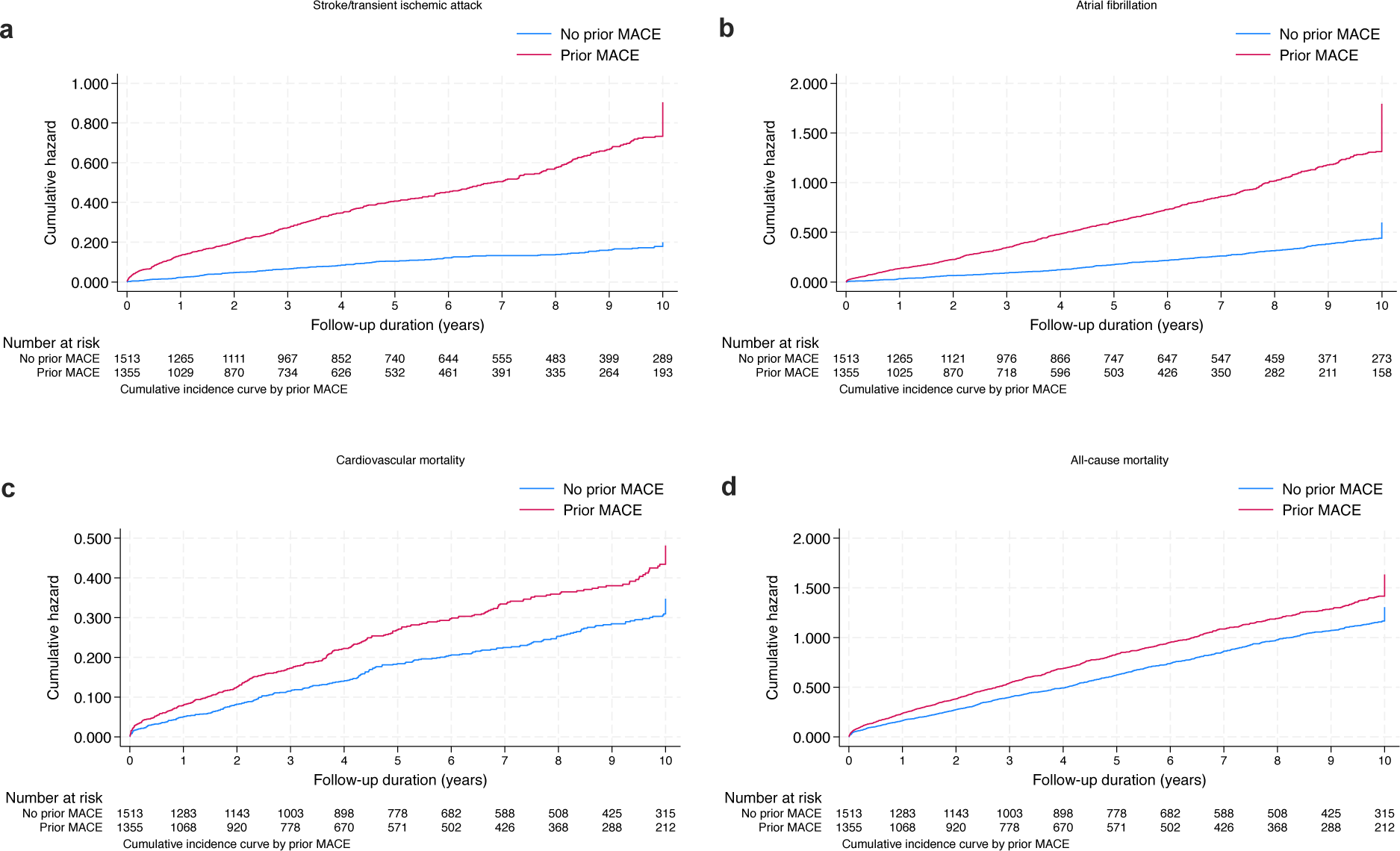
Cumulative incidence curves for primary and secondary outcomes, stratified by prior MACE.

**Fig. 9:**
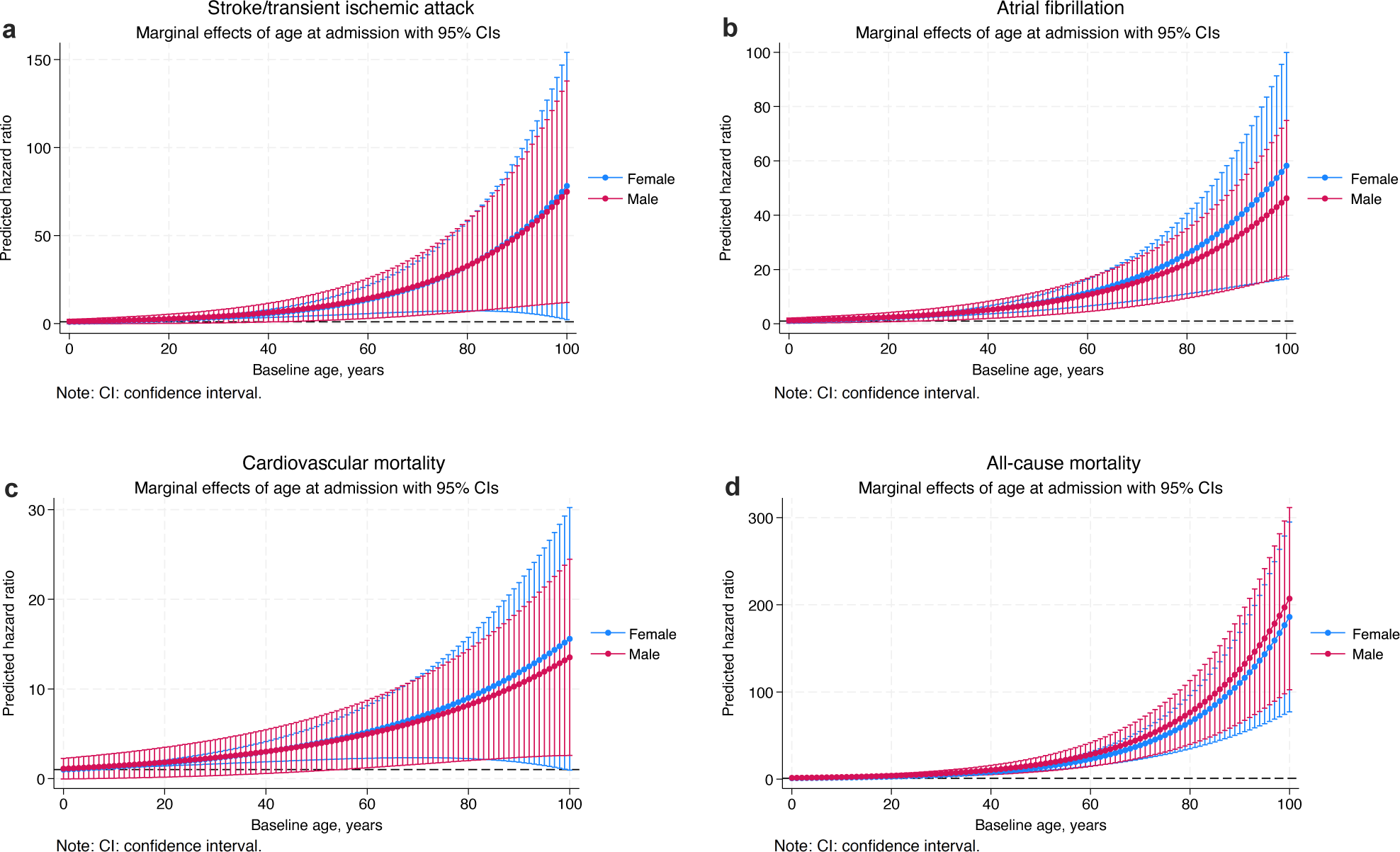
Conditional margin effects analysis of age at admission to predict primary and secondary outcomes in patients with heart failure.

**Fig. 10:**
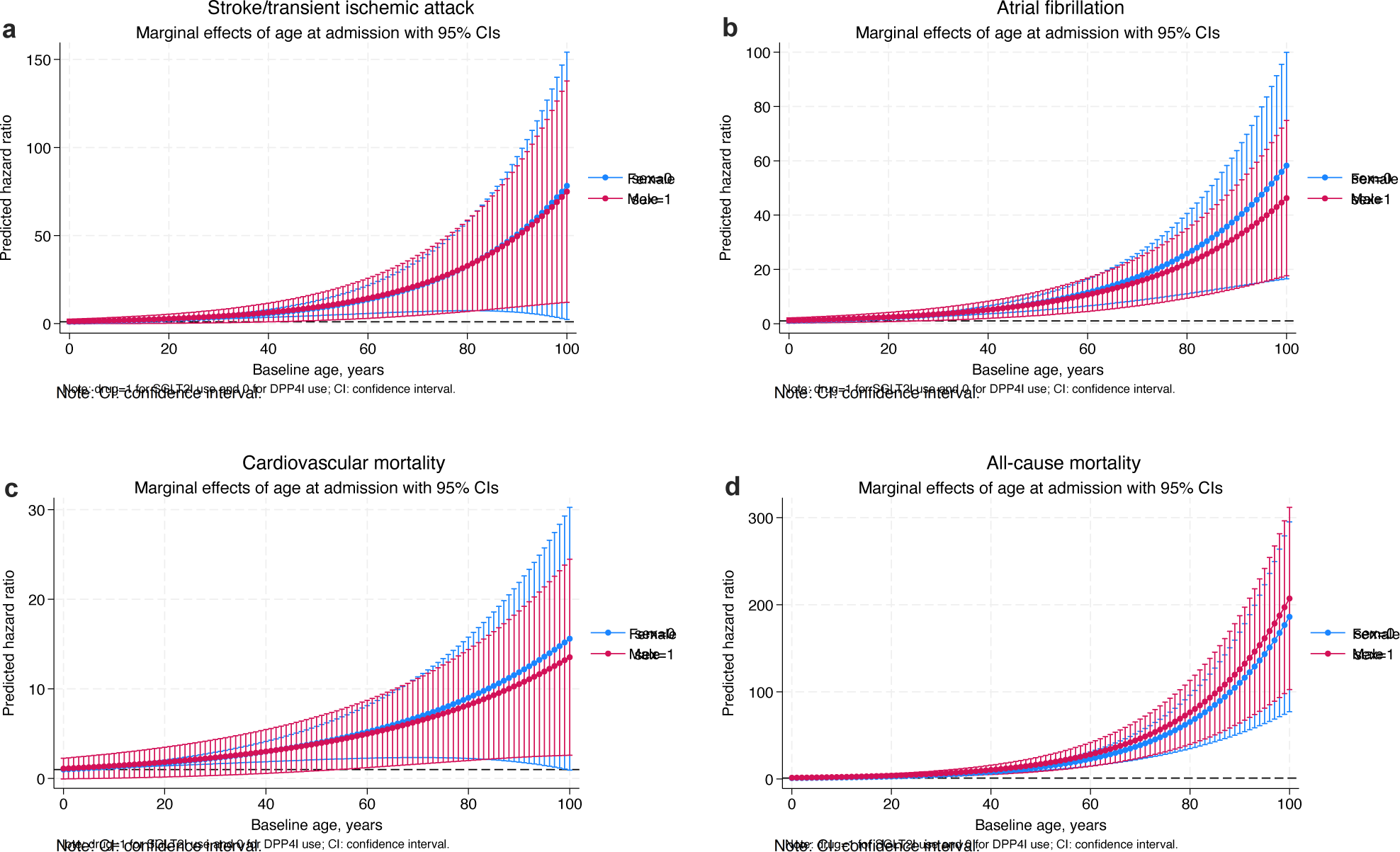
Conditional margin effects analysis of Charlson’s standard comorbidity index to predict primary and secondary outcomes in patients with heart failure.

**Fig. 11:**
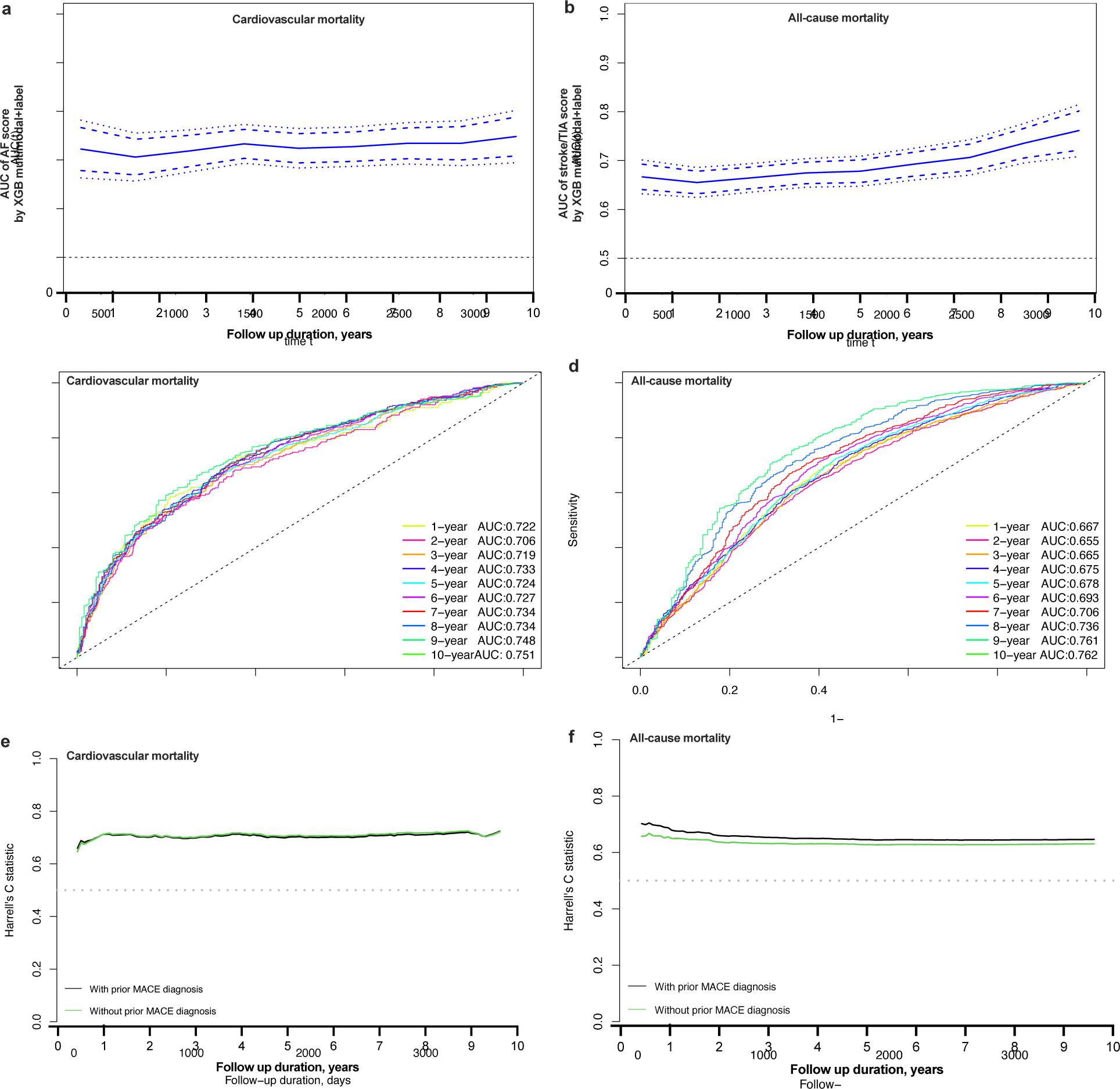
Time-dependent AUROC and Harrell’s C-index of machine learning-based *in-silico* markers to predict mortality risks in heart failure patients. AUROC: Area under the receiver operating characteristic curve. **a-b**. AUROC with 95% confidence interval to predict cardiovascular mortality and all-cause mortality in heart failure patients with the developed *in-silico* marker by XGB multimodal+label model. **c-d**. Prediction performance measured by AUROC to predict cardiovascular mortality and all-cause mortality in heart failure patients with different follow-up duration since admission. **e-f**. Time dependent C-index of the developed *in-silico* marker to predict cardiovascular mortality and all-cause mortality in heart failure patients with different follow-up duration since admission.

## Notes

### Competing Interest Statement

The authors have declared no competing interest.

### Funding Statement

L. L was supported by the InnoHK Project at the Hong Kong Centre for Cerebro-cardiovascular Health Engineering (COCHE). T. Z was supported by the Royal Academy of Engineering under the Research Fellowship scheme.

